# Charting Elimination in the Pandemic: A SARS-CoV-2 Serosurvey of Blood Donors in New Zealand

**DOI:** 10.1101/2021.04.12.21255282

**Authors:** Lauren H. Carlton, Tiffany Chen, Alana L. Whitcombe, Reuben McGregor, Greg Scheurich, Campbell R. Sheen, James M. Dickson, Chris Bullen, Annie Chiang, Daniel J. Exeter, Janine Paynter, Michael G. Baker, Richard Charlewood, Nicole J. Moreland

**Affiliations:** The University of Auckland, Auckland, New Zealand; The New Zealand Blood Service, Auckland, New Zealand; Callaghan Innovation, Christchurch, New Zealand; University of Otago, Wellington, New Zealand

**Author notes:** **Corresponding Author:** Associate Professor Nicole J. Moreland.

**Keywords:** SARS-CoV-2, COVID-19, serosurvey, Spike, receptor binding domain, seroprevalence, New Zealand, elimination

## Abstract

A large-scale SARS-CoV-2 serosurvey of New Zealand blood donors (n=9806) was conducted at the end of 2020. Seroprevalence, after adjusting for test sensitivity and specificity, was very low (0.1%). This finding is consistent with limited community transmission and provides robust evidence to support New Zealand’ s successful elimination strategy for COVID-19.

## Research Letter

New Zealand has a strategy of eliminating SARS-CoV-2 that has resulted in a low incidence of coronavirus-19 disease (COVID-19). The first case was reported on February 26^th^ 2020, and the country entered a strict nationwide lockdown one month later for 49 days (1). Through rigorous border control and managed isolation and quarantine facilities for new arrivals, New Zealand has since remained largely COVID-19 free. Globally, serological surveillance has been utilized throughout the pandemic to define cumulative incidence, including estimations of missed cases and/or asymptomatic infection. Due to lockdowns and movement restrictions, blood donors have been used as a sentinel population in many settings (2,3). The aim of this study was to describe the spread of SARS-CoV-2 in New Zealand via a blood donor serosurvey. Though the pandemic response has been highly effective, PCR testing was initially restricted due to limited diagnostic reagents (4) and there have been occasional border incursions and small community outbreaks, including a cluster in August 2020 with no identified link to the border.

Samples were collected by the New Zealand Blood Service via 9 static collection centers and 36 mobile collection services over a 4-week period (December 3^rd^ 2020 - January 6^th^ 2021) from individuals aged 16 to 88 years. Duplicates were removed, leaving 9,806 samples for analysis. Compared with the 2018 New Zealand census, participants were more likely to be aged 40-59 years (43.3% versus 25.9%) and of European ethnicity (77.8% versus 61.0%) but had a similar proportion of females (49.1% versus 50.7%) and were geographically spread with 16 of 20 district health board regions represented (Table 1 and Appendix). This study was assessed by the Health and Disability Ethics Committee, and additional consent was not required (21/CEN/21).

**Table.**
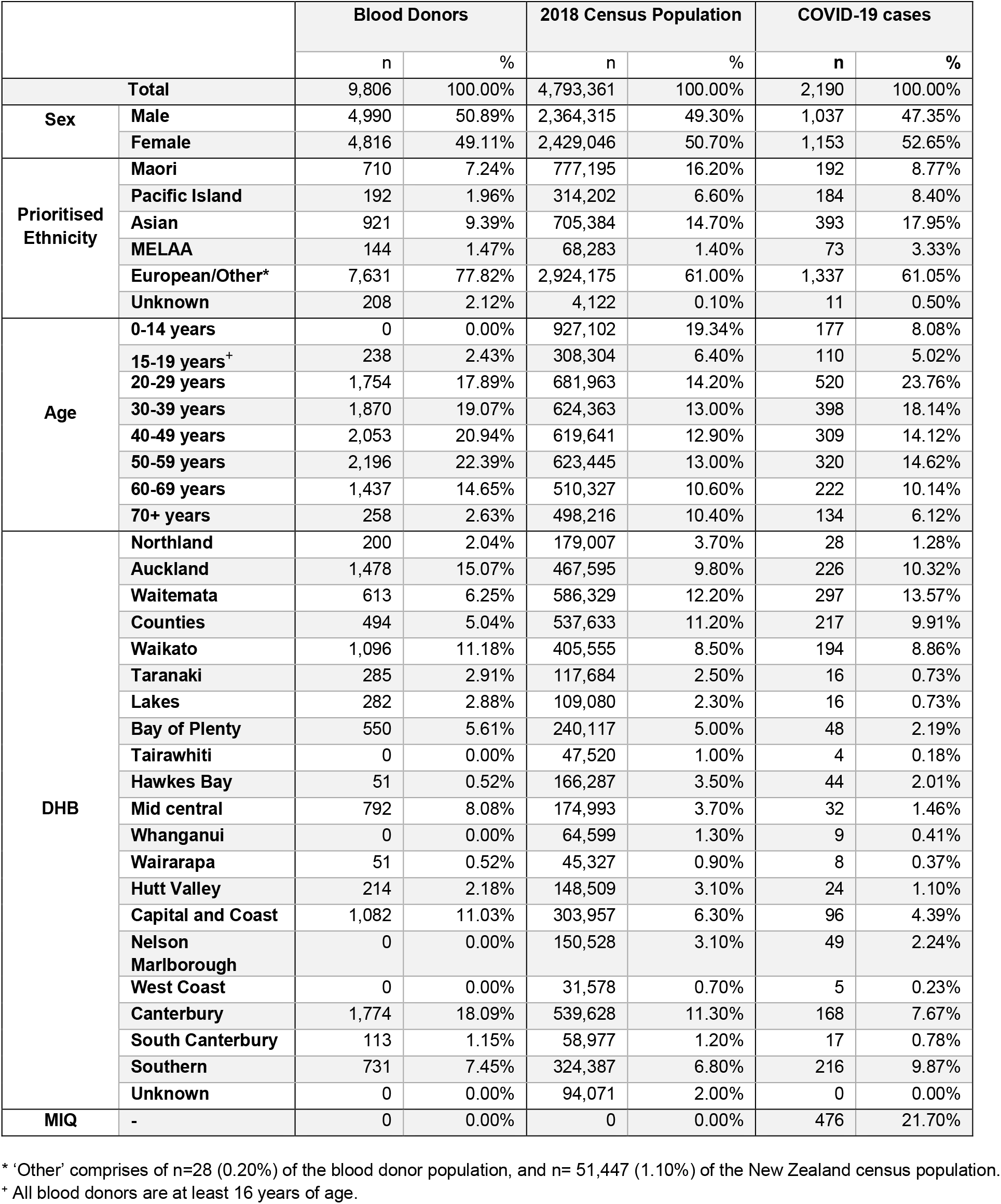
Demographics of the blood donors, 2018 Census population and COVID-19 cases in New Zealand. The New Zealand blood service donations were collected between the 3^rd^ of December 2020 and the 6^th^ of January 2021. Of the 9,806 individuals, 9,771 are blood donors and 35 are living tissue and stem cell donors. All donors must be free of illness and weigh >50 kg. Notable travel and a history SARS-CoV-2 infection (or contact with a positive case) are recorded prior to collection. Demographics for COVID-19 cases were obtained from the New Zealand Ministry of Health and includes probable and confirmed infections up to and including the 6^th^ of January 2021. The most recent New Zealand census took place in March 2018. Priority ethnicity is reported as defined by the New Zealand Department of Statistics. Abbreviations: MELAA, Middle Eastern, Latin American and African; DHB, District Health Board; MIQ, Managed Isolation and Quarantine.

Antibodies to the Spike (S) protein and receptor-binding domain (RBD) persist for many months after infection, compared with antibodies to the nucleocapsid (N) protein (5,6), providing rationale for the use of S protein-based assays in serosurveys. The overall serological testing algorithm was optimized for specificity given the low number of reported COVID-19 cases in New Zealand (2,190 as of January 6^th^ 2021) and the associated period prevalence of 0.04%, which limits the positive predictive value of tests with reduced specificity (7). Samples were first screened with a widely used and well-validated 2-step ELISA that comprises a single point dilution assay against the RBD followed by titration against trimeric S protein (Appendix) (8,9). Samples above the cut-off were tested on two further immunoassays – the EuroImmun SARS-CoV-2 IgG ELISA (EuroImmun AG, Lübeck, Germany) and the cPass surrogate Viral Neutralization Test (sVNT) (GenScript, New Jersey, USA) and deemed seropositive if above the cut-off on both commercial assays. Sensitivity and specificity for these assays were determined by Receiver Operator Characteristic (ROC) curves based on previous analyses (413 pre-pandemic negatives, 99 PCR confirmed cases) (Appendix) (9,10).

Of the 9,806 samples, 18 were positive for both Spike IgG (EuroImmun) and antibodies that block the RBD-hACE-2 interaction (sVNT), with the values highly correlated (Pearson r 0.7993, p < 0.0001) (Figure). Further analysis of the 18 seropositive samples with a multiplex bead-based assay that detects antibody isotype reactivity to RBD, S and N proteins (5) revealed a pattern consistent with infections that occurred weeks or months prior; a dominance of RBD and S protein IgG with few samples positive for N protein IgG, nor IgA or IgM against any of the three antigens (Figure). Within these 18 seropositive samples, six were retrospectively matched to donors with previously confirmed SARS-CoV-2 infections. That all confirmed cases were detected supports the rationale of the testing algorithm applied. A further four seropositive samples were from donors with 2020 travel history in settings with a high risk of SARS-CoV-2 exposure (United Kingdom and Europe), suggesting likely infection outside New Zealand. The remaining eight seropositive samples were from seven different district health regions, giving a crude seroprevalence estimate of 0.082% (95% confidence intervals [CI] 0.035-0.16%). Applying the Rogan-Gladen estimate with the Lang-Reiczigel CI method to account for test sensitivity and specificity results in a true seroprevalence estimate of 0.103% (95% CI 0.09-0.12%) (Appendix) and an infection to case ratio of 2.3, based on notified cases on January 6^th^, 2021, suggesting some undiagnosed infections have occurred. However, this ratio needs to be interpreted with caution given the limitations of the sampling population, together with the extremely low number of seropositive donors, which also precludes any subgroup analysis.

**Figure.**
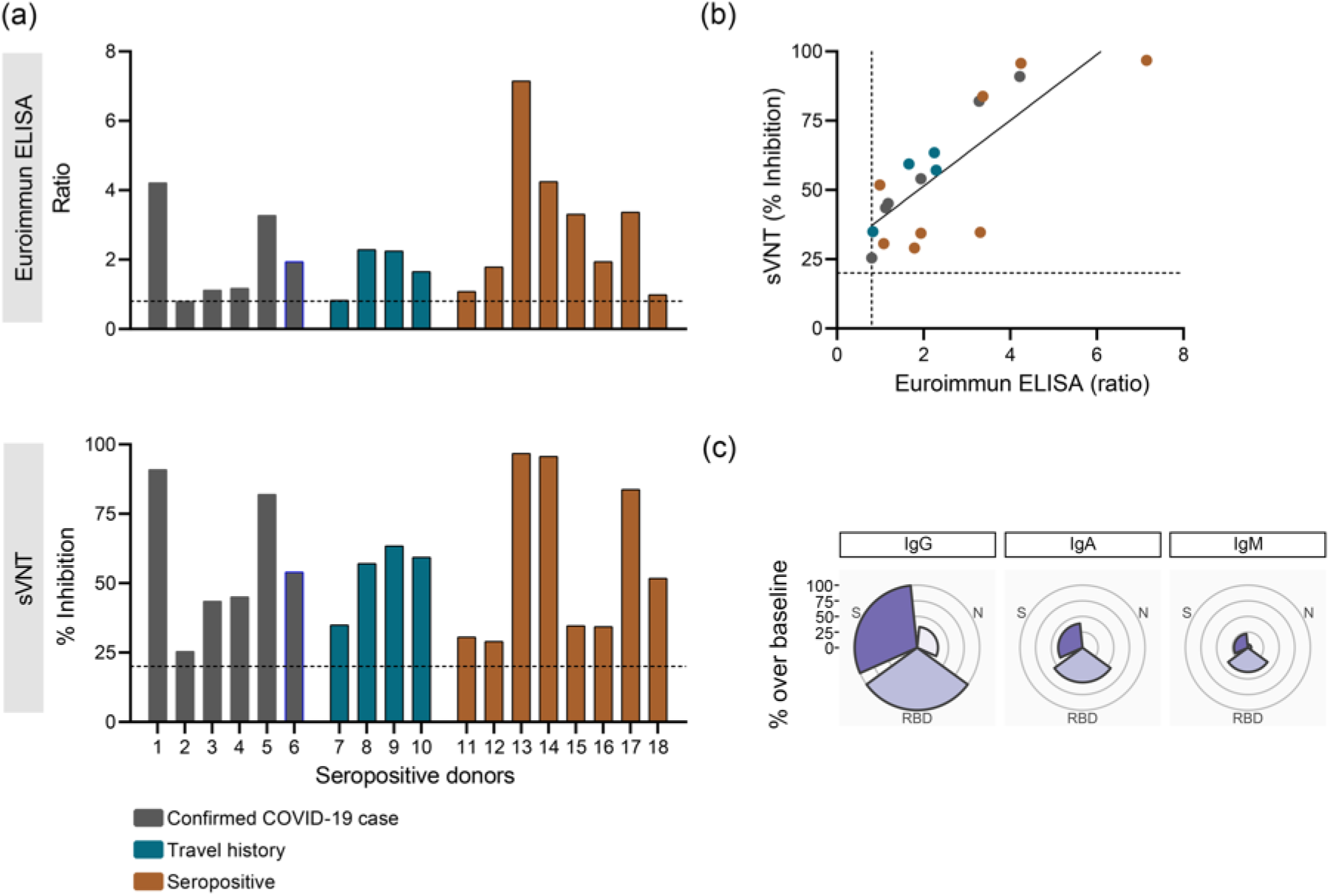
Antibody characteristics of the seropositive donors (n=18). (a) Seropositivity was confirmed by EuroImmun S1 IgG (top) and the surrogate Viral Neutralization Test (sVNT, bottom). Six donors had PCR confirmed SARS-CoV-2 infection (dark grey), four had relevant travel history (dark turquoise), and eight were identified in this study (orange). The manufacturers cut-offs are shown (black dotted line). (b) Pearson correlation of sVNT and the Euroimmun IgG ELISA (n=18). (c) Rose plot showing percentage of seropositive donors over baseline for IgG, IgA and IgM antibodies against the RBD, Spike (S) and Nucleocapsid (N) proteins determine using a multi-plex Luminex bead assay.

The very low seroprevalence of SARS-CoV-2 infection in New Zealand implies undetected community transmission has been limited. This seroprevalence is broadly similar to a recent study conducted in a low prevalence state of Australia (3), and markedly lower than estimates of >10% from serosurveys in Europe and America where the pandemic has been poorly controlled (https://serotracker.com). This study provides the first robust, serological evidence of New Zealand’ s successful elimination strategy ahead of vaccine roll-out.

## Data Availability

The data that support the findings of this study are available on request from the corresponding author, NJM, upon reasonable request.

## Acknowledgments

This work was funded by the School of Medicine Foundation (University of Auckland), and the COVID-19 Innovation Acceleration Fund (Ministry of Business, Innovation and Employment). The 2018 Census data used in this study were supplied by Statistics New Zealand (Stats NZ) and accessed via its Integrated Data Infrastructure (IDI).

## Statistics New Zealand Disclaimer

Access to the data used in this study was provided by Stats NZ under conditions designed to give effect to the security and confidentiality provisions of the Statistics Act 1975. The results presented in this study are the work of the author, not Stats NZ or individual data suppliers.

