## Supplementary Material for "Charting Elimination in the Pandemic: A SARS-CoV-2 Serosurvey of Blood Donors in New Zealand"

### Appendix

#### Blood donor participants

The sample population is made up of 9,806 individuals (9,771 blood donors and 35 living tissue and stem cell donors) who had donated to the New Zealand Blood Service between December 3rd, 2020 and January 6th, 2021. The minimum donor age is 16, and minimum weight is 50 kg. Standard donor exclusions include illness within the previous 28 days as well as vCJD risk if domiciled in the UK between 1980-1996. Notable travel and history of a SARS-CoV-2 infection (or contact with a positive case) are recorded prior to collection. Donors are deferred for 28 days after the last known contact with a person with confirmed SARS-CoV-2 infection or resolution of all COVID-19 symptoms. The type of collection centre and donation type are shown (Supplementary Table 1). The age range for blood donation is 16-76 years, with no upper limits for tissue and haematopoietic stem cell donation. The sample population is geographically spread through New Zealand, with sixteen of twenty New Zealand Health Board regions represented (Supplementary Figure 1). The most populous regions based on the 2018 National census are well covered by the survey population, while the regions not represented (Tairawhiti, Whanganui, Nelson Marlborough and the West Coast) comprise lower proportions of the New Zealand population (1-2 or 3-4%).

**Supplementary Table 1.** Donation venue types for study participants; HPC: Hematopoietic Progenitor Cell

| Venue Type | No. of venues | % donors |
|---|---|---|
| **HPC** | 3 | 0.1% |
| **Mobile** | 36 | 25.5% |
| **Static** | 9 | 74.1% |
| **Surgical** | 19 | 0.3% |
| **Total** | **67** | **100%** |

**Supplementary Figure 1.** A map representation of the New Zealand blood donor serosurvey participants compared with the New Zealand population at the most recent census conducted in 2018. Percentage representation of each region is shown as a colour scale from white to dark red (blood donors) or dark green (2018 census). Maps were created using ArcGIS version 10.7.1

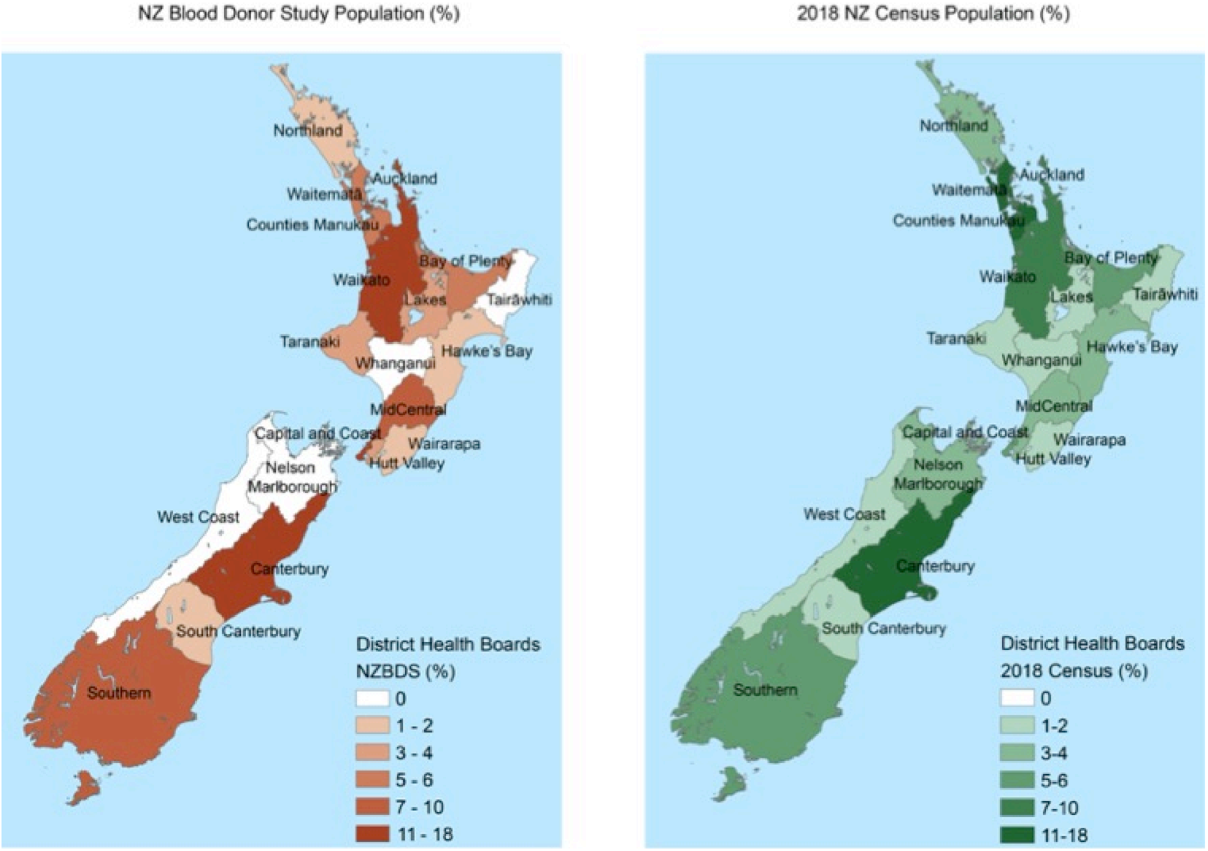

#### SARS-CoV-2 Serologic Assay Methodology

*In-house two-step ELISA*

The *in-house* two-step ELISA and was performed as previously described using Spike (S) protein and receptor binding domain (RBD) antigens produced in a 293 human embryonic kidney cell expression system (*1*,*2*). In the first step, serum (1:100) was screened against RBD coated immunoplates (5 μg/ml), with IgG binding detected using a peroxidase-labelled anti-human IgG secondary (97221; Abcam, Cambridge, United Kingdom). Samples with an optical density (OD, 450-570nm) above the cut-off (OD >0.2) in the RBD ELISA were titrated in a 3-fold dilution series starting at 1:100 dilution against S protein coated immunoplates (5 μg/ml) and considered positive if the OD was >0.2 in at least two consecutive wells. Positive and negative controls were included on each plate, with the assay meeting acceptance criteria if the OD was >0.6 and <0.02 for the positive and negative control, respectively. The positive control comprised a pool of PCR positive sera from the AccuSet SARS-CoV-2 Performance Panel, while the negative control was PCR negative serum from the same AccuSet Panel (Seracare, Massachusetts, USA). To track assay stability, additional positive and negative quality controls (Virotrol (positive) and Viroclear (negative), Biorad, California US) were included on each plate (Supplementary Figure 2).

**Supplementary Figure 2.** Quality control variance for the *in-house* 2-step ELISA. The SARS-CoV-2 positive (a) and negative (b) sera from (Biorad, California US) were included on all ELISA plates. Data is shown for quality controls included on each RBD ELISA plate (n=113) over an 8-week period as a weekly average. Levey Jennings analyses shows the values are within mean (black line) and two standard deviations (2-SD) with 1-SD (light blue), 2-SD (mid-blue) and 3-SD (dark blue) as shown. The inter-plate coefficient of variance (CV) for the positive quality control was 9.2% across all plates.

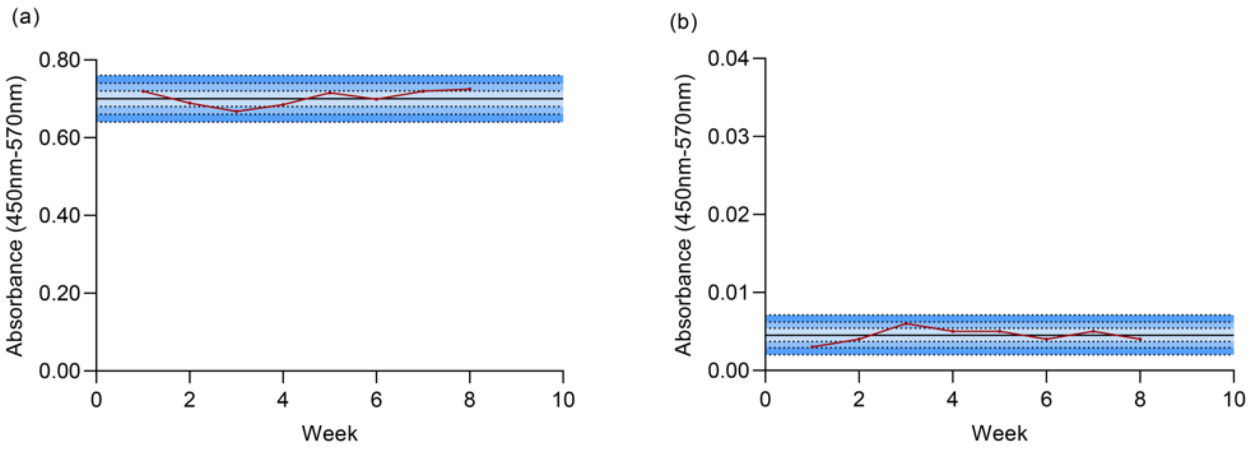


*Euroimmun S1 IgG ELISA and Surrogate Viral Neutralisation assay (sVNT)*

The EuroImmun SARS-CoV-2 ELISA (Euroimmun, AG Lübeck, Germany) detects IgG antibodies targeting the S1 domain of the SARS-CoV-2 S protein and was carried out according to manufacturer's instructions. A ratio was calculated of the OD (450-650nm) of the sample divided by the OD of the supplied calibrator and samples with a ratio of ≥0.8 were considered positive. The cPass® surrogate viral neutralisation test (sVNT) (GenScript, New Jersey, USA) measures antibodies (NAbs) that block the interaction between RBD and hACE2 (*3*) and was performed according to manufacturer's instructions. Samples with percentage inhibition ≥20%

were considered positive. The sensitivity and specificity for these assays were determined by Receiver Operator Characteristic (ROC) curves based on previous analyses (413 pre-pandemic negatives, 99 PCR confirmed cases) *(2,4)*. The Euroimmun S1 IgG ELISA has a sensitivity of 91.92% (95% confidence interval (CI): 84.86 to 95.85%) and specificity of 98.79% (95% CI: 97.20 to 99.48%) at the manufacturers recommended cut-off of ratio of 0.8. The sVNT has a sensitivity of 88.89% (95% CI: 81.19 to 93.68%) and a specificity of 100.00% (95% CI: 99.08 to 100%) at the manufacturers recommended cut-off of 20% inhibition (Supplementary Figure 3).

**Supplementary Figure 3.** Receiver operating characteristic (ROC) curves for the two commercial assays used to confirm seropositivity. The line of no discrimination is shown in red.

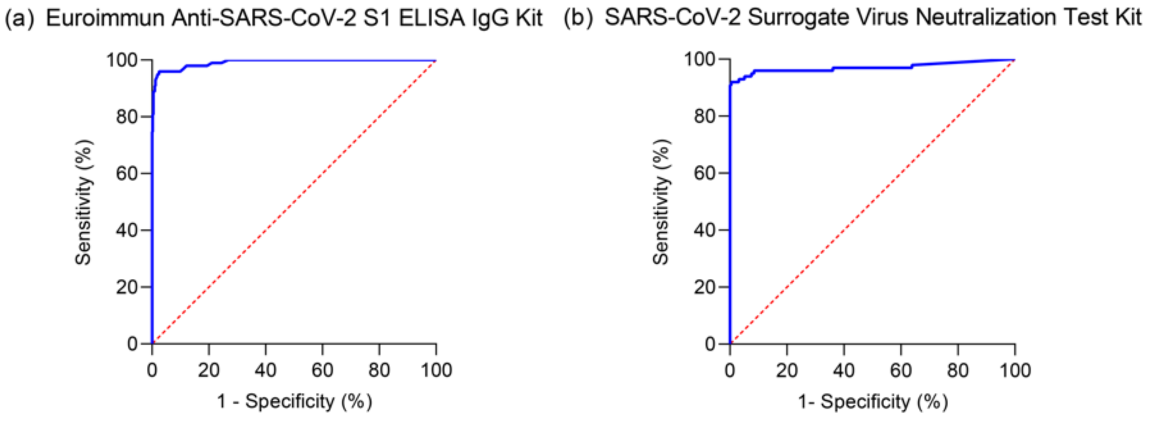


*Multiplex bead-based assay*

The luminex multiplex bead-based assays used to detect antibody isotype reactivities to viral proteins were conducted as previously described (*5*). Briefly, samples were diluted to 1:400 (IgA, IgM) or 1:800 (IgG) in assay buffer and incubated with RBD, S and N protein beads.

PE- labelled anti-human IgA/M/G (Southern Biotech, Alabama, US) were used to detect the different isotypes.

**Prevalence estimations**

The 95% confidence intervals of the crude seroprevalence were estimated using the conservative exact Clopper-Pearson method (*6*). For true prevalence calculations, a combined sensitivity of 0.8171 and specificity of 1 was used as the two tests were used in series, i.e. samples needed to test positive on both commercial assays to be considered positive. This method increases specificity at the expense of sensitivity. As true prevalence was expected to be close to 0 it was estimated using the Rogan-Gladen point estimate for prevalence *(7*) with the Lang-Reicziegel confidence interval method (*8*). This adjusts for the fact that test sensitivity is estimated from a sample of true positive individuals (n=99) and the specificity is an estimate from a sample of true negative individuals (n=413). The method has been shown to provide good coverage for prevalence close to 0 and overcomes several short comings of traditional methods *(8,9).*
